# Routes for COVID-19 importation in Brazil

**DOI:** 10.1101/2020.03.15.20036392

**Authors:** Darlan Da S Candido, Alexander Watts, Leandro Abade, Moritz UG Kraemer, Oliver G Pybus, Julio Croda, Wanderson de Oliveira, Kamran Khan, Ester C Sabino, Nuno R Faria

## Abstract

**Highlight:** The global outbreak caused by the severe acute respiratory syndrome coronavirus-2 (SARS-CoV-2) has been declared a pandemic by the WHO. As the number of imported SARS-CoV-2 cases is on the rise in Brazil, we use incidence and historical air travel data to estimate the most important routes of importation into the country.

## Main Text

Severe acute respiratory syndrome coronavirus-2 (SARS-CoV2) was first detected in Wuhan, Hubei province, China, on December 8^th^ 2019. SARS-CoV-2 infection can cause coronavirus disease (COVID-19) and can lead to acute respiratory syndrome, hospitalization and death.^1^ As of the 12^th^ March 2020, the global SARS-CoV-2 outbreak has been declared a pandemic, with 125,048 cases and 4,613 deaths have been notified by the World Health Organization (WHO) in 117 countries/territories or areas worldwide (who.int/emergencies/diseases/novel-coronavirus-2019/situation-reports). The first case in Latin America was confirmed on February 26, 2020, in the São Paulo metropolis, the most populous city in the Southern hemisphere (∼11 million people Instituto Brasileiro de Geografia e Estatística, www.ibge.gov.br). Self-declared travel history and subsequent genetic analyses confirmed that this infection was acquired via importation of the virus from Northern Italy^2^. Since then Brazil has reported the largest number of cases in Latin America (n=34, as of March 10, 2020). SARS-CoV-2 has been now detected in 7 (26%) of the 27 federal states of Brazil. So far, transmission of SARS-CoV-2 appears to be primarily sporadic (85.3%, 29/34 are imported cases). Here, we analyse data on airline travellers to Brazil in 2019, who departed from countries that had reported local cases of COVID-19 transmission by March 5^th^ 2020. This information provides insights into which Brazilian cities are most at risk for SARS-CoV-2 importation.

We used travel data on all air journeys that had a Brazilian city as their final destination during February and March 2019 as a proxy for flight density during the 2020 COVID-2019 outbreak (see Supplementary Material). We focused on the data for 29 countries that had reported SARS-CoV-2 cases by 5^th^ March 2020. We collated the total number of passengers flying to any Brazilian airport during this period, country population size for 2019 from the United Nations World Population Prospects 2019 database, and the WHO-reported number of COVID-19 cases (as of March 5^th^, 2020). We used these values to estimate the proportion of infected travellers potentially arriving in Brazilian cities from each country and for each route (additional information can be found in Supplementary Material). No air passenger data from Iran to Brazil was available for our analysis.

Between February and March 2019, Brazil received 841,302 international passengers in a total of 84 cities across the country (Figure 1). São Paulo, the largest city in the country, was the final destination of nearly half (46.1%) of the passengers arriving to Brazil, followed by Rio de Janeiro (21%) and Belo Horizonte (4.1%). More than half of the international passengers started they journey in the USA (50.8%) followed by France (7.9%) and Italy (7.5%). The air-travel routes to airports in Brazil with most passengers were USA-São Paulo (23.3%), USA-Rio de Janeiro (9.8%) and Italy-São Paulo (3.4%).

**Figure 1.**
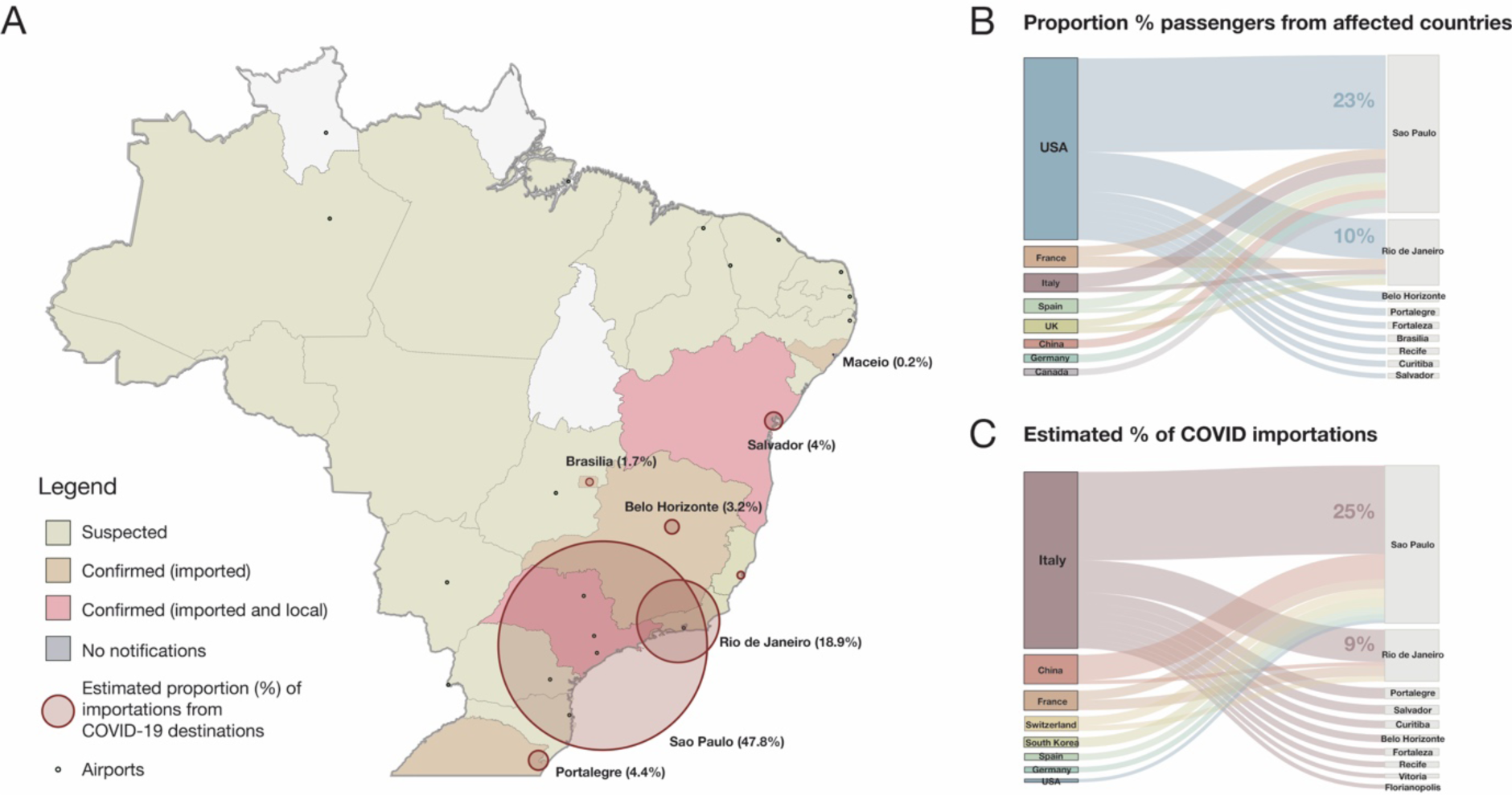
Potential for COVID-19 importation to Brazil. A) Map of Brazilian federal states and Federal District coloured according to COVID-19 notification status (as of March 10, 2020). Circles correspond to the estimated proportion of arrivals from the top 29 destinations (except Iran) that had reported local COVID-19 by 5^th^ March 2020. B) Percentage of passengers for the top-20 routes to Brazilian airports from countries that had reported COVID-19 cases by the 5^th^ March 2020. C) Estimated percentage of importations for the top-20 routes from countries that had reported local COVID-19 by the 5^th^ March 2020.

To better understand the potential for SARS-CoV-2 introductions to Brazil, we estimate the relative risk of COVID-19 introduction to Brazilian cities by taking into account SARS-CoV-2 incidence per international traveller arriving at an airport in Brazil. We estimate that 54.8% of all imported cases would be expected to come from travellers infected in Italy, 9.3% and 8.3% of the cases would be from travellers infected in China and France, respectively. The route Italy-São Paulo was estimated to comprise 24.9% of total infected travellers travelling to Brazil during this period. Moreover, we estimate that Italy has been the source location for five of the top 10 most importation routes for infected travellers into Brazil based on the current epidemiological scenario (Supplementary Information). Consistent with this, at least 48% (n=14/29) of the reported imported cases in Brazil have a history of travelling to Italy prior to onset of symptoms, as of 9^th^ March 2020. Six (23.1%) of the confirmed cases that acquired the virus in Italy have been identified in São Paulo (Supplementary Information).

We find that the proportion of estimated imported cases by airport of destination is highly correlated with the proportion of detected imported cases. Our study has several limitations. Unfortunately, data from Iran was not available for this analysis. Moreover, our analysis relies on incidence data, and thus the risk of importation will follow changes in epidemic sizes at source locations. In fact, with the reduction in the number of flights leaving from Italy and 51% of flights to Brazil depart from airports in the USA, we should anticipate for an increasing proportion of infected travellers arriving from the USA. Moreover, the estimated risk of importation from China is likely an overestimate as recent measures have extensively decreased the flights to Brazil.

At a time when the number of SARS-CoV-2 cases are steadily growing in Brazil, our findings highlight the high potential for the introduction of new cases in several cities of Brazil, especially in Sao Paulo and Rio de Janeiro metropolises. Rapid identification of locations where clusters of local transmission might first ignite is critical to better coordinate preparedness, readiness and response actions.^3,4^ There is critical need for epidemiological, human mobility and genetic data^5^ to understand virus transmission dynamics at local, regional and global scales. Continued integration of these data streams should help guide deployment of resources to mitigate COVID-19 transmission.

## Data Availability

Data will be made available upon request.

## Authors Statements

KK is the founder of BlueDot, a social enterprise that develops digital technologies for public health. KK and AW are employed at BlueDot. DSC, LA, MK, WO, JC, ECS, OGP, NRF have no conflicts of interest to declare.

## Authors Contributions

DSC, LA, NRF conceived the idea and wrote the manuscript. DSC, LA, NRF, KK, AW conducted data analysis. DSC, NRF, LA, MUGK, WO, JC, ECS, OGP, AW, KK interpreted data and contributed to writing.

